# Improving Privacy and Utility in Aggregate Data: A Hybrid Approach

**DOI:** 10.1101/2024.05.05.24306903

**Authors:** Samuel Nartey Kofie, Ivy Min-Zhang, Kai Chen, Wei Percy

## Abstract

The increasing need to protect individual privacy in data releases has led to significant advancements in privacy-preserving technologies. Differential Privacy (DP) offers robust privacy guarantees but often at the expense of data utility. On the other hand, data pooling, while improving utility, lacks formal privacy assurances. Our study introduces a novel hybrid method, termed PoolDiv, which combines differential privacy with data pooling to enhance both privacy guarantees and data utility. Through extensive simulations and real data analysis, we assess the performance of synthetic datasets generated via traditional DP methods, data pooling, and our proposed PoolDiv method, demonstrating the advantages of our hybrid approach in maintaining data utility while ensuring privacy.

## 1 Introduction

Privacy-preserving synthetic data generation and analysis has gained considerable attention in various fields ranging from heath care to social media [1, 2]. The recent surge in the amount of data collected, in health-care for example, provides interesting avenues for exploration and insightful discovery. However, collecting and sharing sensitive, usable, micro-data without disclosing personal information is challenging. In some cases, e.g. through adversarial attacks such as linkage reidentification of individual participants is a possibility.[3]

Differential privacy provides a formal guarantee for confidentially [4, 5]. The mechanism promises that an individual participant’s record would remain private even if the adversary has complete knowledge of the rest of the database. This stringent promise has made DP the gold standard for data release [6]. Most agencies provide differentially private synthetic datasets which are often labeled as true representations of the sensitive dataset. [7] Consequently, users must implicitly assume the agency (or curator) has a detailed knowledge of the data characteristics, and that the synthetic data generation model employed by the DP mechanism is well defined. In many cases, users are unable to determine if and how much their analysis results have been impacted by the synthesis process. Inevitably, the accuracy of some analyses deteriorates significantly due to imperfect data generation models. Another formal complain DP synthetic data, is that the mechanism destroys potential insightful data structure such as voids or manifolds.

Another synthetic data generation method that has gained prominence over the years is data aggregation (also referred to as specimen pooling) [6, 8, 9]. Pooling offers an attractive alternative to the differential privacy mechanism as it is much easier to implement in practice. However, it doesn’t provide a formal privacy guarantee. The technique randomly combines information from individuals of the same outcome category or exposure of interest, which is then shared with an analyst. This method suggests preserving privacy by sharing aggregate data instead of individual-level data.[10, 11, 12]

The present study was structured around three primary objectives:

**1. Performance Comparison:** Our first goal was to assess the performance of synthetic data generated from traditional Differential Privacy (DP) mechanisms in comparison to those generated through pooling mechanisms. This evaluation utilized regression modeling to analyze the impact of each method on data utility and accuracy.

**2. Hybrid Mechanism Development:** We proposed a novel hybrid mechanism that combines differential privacy with data pooling, referred to as the pooled-DP mechanism. This initiative was undertaken to enhance privacy protection in pooled data setups without compromising data utility. Subsequently, the performance of this hybrid mechanism was compared with traditional DP and pooling methods to ascertain its effectiveness.

**3. Data Clustering Analysis:** The third objective focused on examining the clustering patterns of synthetic data produced using both the newly developed pooled-DP mechanism and traditional methods. Analyzing these patterns aids in understanding how various synthetic data generation techniques influence the underlying structures and relationships within the data.

These objectives were designed to collectively advance our understanding and refinement of data privacy techniques, ensuring robust privacy protections while maintaining the synthetic data’s utility and quality for analytical purposes.

## 2 Related Work

This section explores the foundational techniques employed in the generation of synthetic data, particularly focusing on ensuring confidentiality and utility. Differential privacy and data pooling methods have been extensively used to address these dual objectives. Here, we delve into differential privacy, its definitions, key mechanisms, and their application in data protection.

### 2.1 Differential Privacy

Differential Privacy (DP) was introduced as a robust framework for protecting the confidentiality of individual data in datasets used for research and analysis [5]. It is designed to offer strong protection against adversaries who may have access to auxiliary information, thus ensuring that the participation of any individual in a dataset does not significantly influence the outcome of any analysis. For a survey of differential privacy, we refer reader to these articles [13, 14].

*Definition 1 (ϵ-Differential Privacy):* A mechanism ℳ is said to satisfy *ϵ*-differential privacy if for any two adjacent datasets *D* and *D*^′^ that differ by a single individual’s data, and for all events *Z* in the output space of ℳ, the probability that ℳ outputs *Z* satisfies the following inequality:

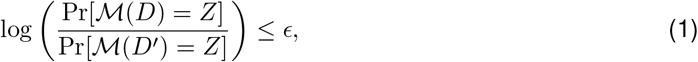

where *ϵ >* 0 is a small constant that determines the level of privacy. The smaller the *ϵ*, the greater the privacy protection, as the outputs from *D* and *D*^′^ are made statistically more indistinguishable.

The definition can be extended to (*ϵ, δ*)-differential privacy to allow a small probability *δ* of the mechanism failing to meet the *ϵ*-differential privacy condition. This relaxation is particularly useful when dealing with complex data or when aiming to improve the utility of the data after applying privacy-preserving techniques.

Within this framework, several mechanisms have been developed to enforce differential privacy:

*1. Randomized Response Mechanism*: This mechanism enhances privacy by introducing randomness in the responses. For example, in a survey, respondents might flip a biased coin in private and only provide their true answer if the coin comes up heads. This simple approach provides a foundational layer of privacy by dissociating the individual’s response from their actual data.

*2. Laplace Mechanism*: One of the most common methods for implementing differential privacy involves adding noise generated from a Laplace distribution to the query results. The scale of the Laplace noise is proportional to the sensitivity of the function being computed over the data (denoted as 𝕊 _ℳ_) and inversely proportional to *ϵ*, ensuring that the added noise maintains the utility of the data while protecting individual privacy.

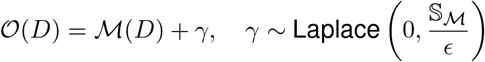

*3. Exponential Mechanism*: Used primarily for selecting outputs from a set of possible outcomes. This mechanism assigns probabilities to these outcomes based on a scoring function, which quantifies the utility of each outcome. The selection is skewed towards outcomes with higher utility, adjusted exponentially in accordance with the privacy parameter *ϵ*.

*Definition 2 (Sensitivity):* The sensitivity of a query function ℳ, essential for calculating the requisite noise addition in differential privacy, is defined as the maximum change in the output of ℳ when any single individual’s data is changed or removed.

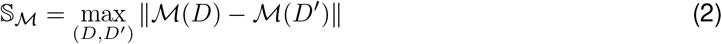

swhere ∥ · ∥ denotes the L_1_ norm. The effectiveness of these mechanisms is illustrated in Appendix A (Figure **??**), showing how different privacy budgets impact the degree of noise and hence the privacy-utility trade-off.

#### 2.1.1 *ϵ*-Differential Private Synthetic Data

Differential privacy has emerged as a robust framework for data sharing, particularly attractive for its stringent privacy guarantees. This framework has spurred the development of both interactive and non-interactive methods for data release, which adapt various techniques to balance privacy with data utility [15, 16].

We focus on two primary categories of differential privacy mechanisms for generating synthetic data: non-parametric and parametric methods.

##### Non-parametric Methods

Non-parametric methods do not assume any underlying statistical model for data generation. Instead, they directly utilize the empirical distributions of data attributes to generate synthetic data [5]. One common approach is the histogram perturbation method, where noise is added to the histograms of the data attributes to protect privacy before generating synthetic data from the perturbed histograms.

###### Algorithm 1

Histogram Perturbation Method

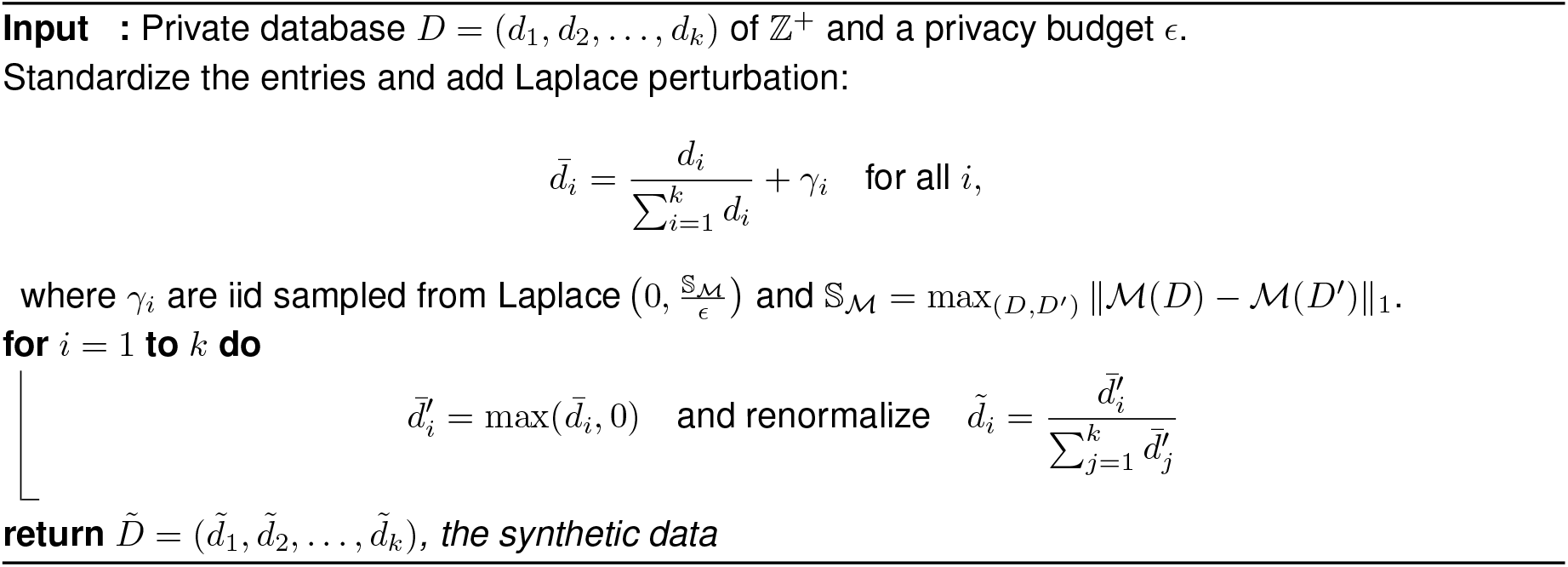

##### Parametric Methods

Parametric methods assume a statistical model for the original data and use parameters estimated from the data to generate synthetic datasets. These methods often use a Bayesian approach, where data is generated from a distribution defined by posterior parameters perturbed according to differential privacy requirements.

###### Algorithm 2

Multinomial-Dirichlet Synthetic Data Generation

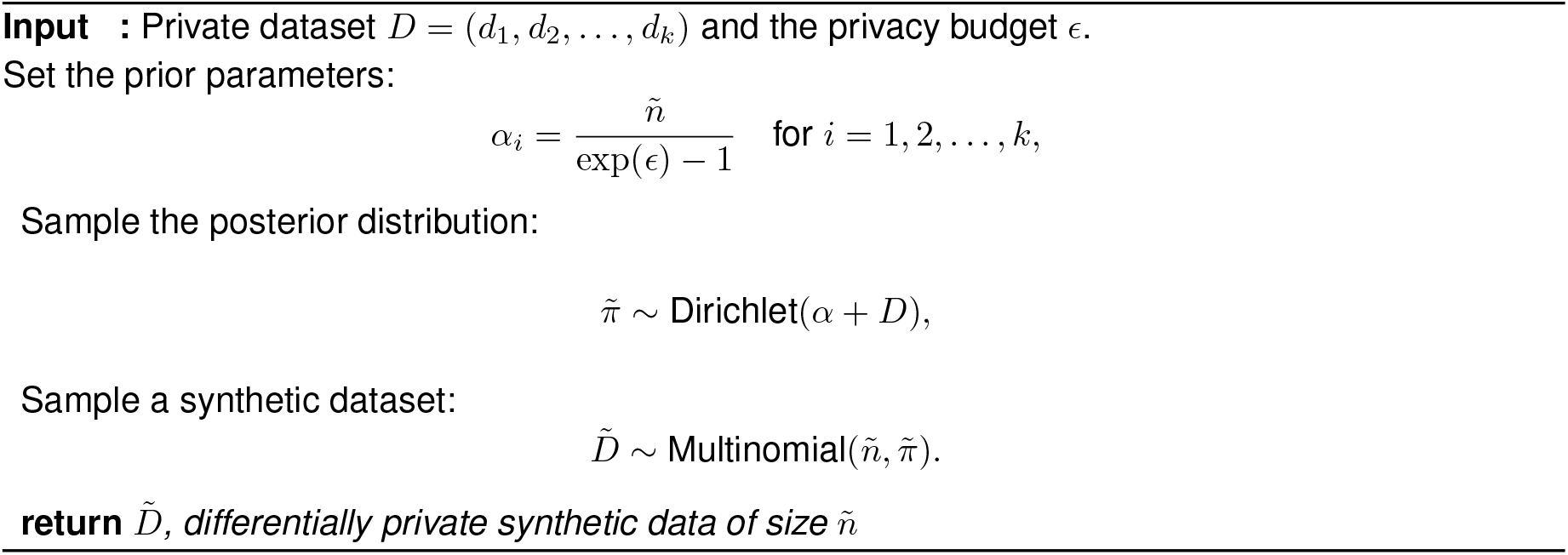

Figure 1 presents a schematic representation of these mechanisms, illustrating the general approach to generating synthetic data under differential privacy. Both methods are vital in scenarios where the original data must remain confidential, yet the utility of the data for analysis cannot be compromised.

**Figure 1:**
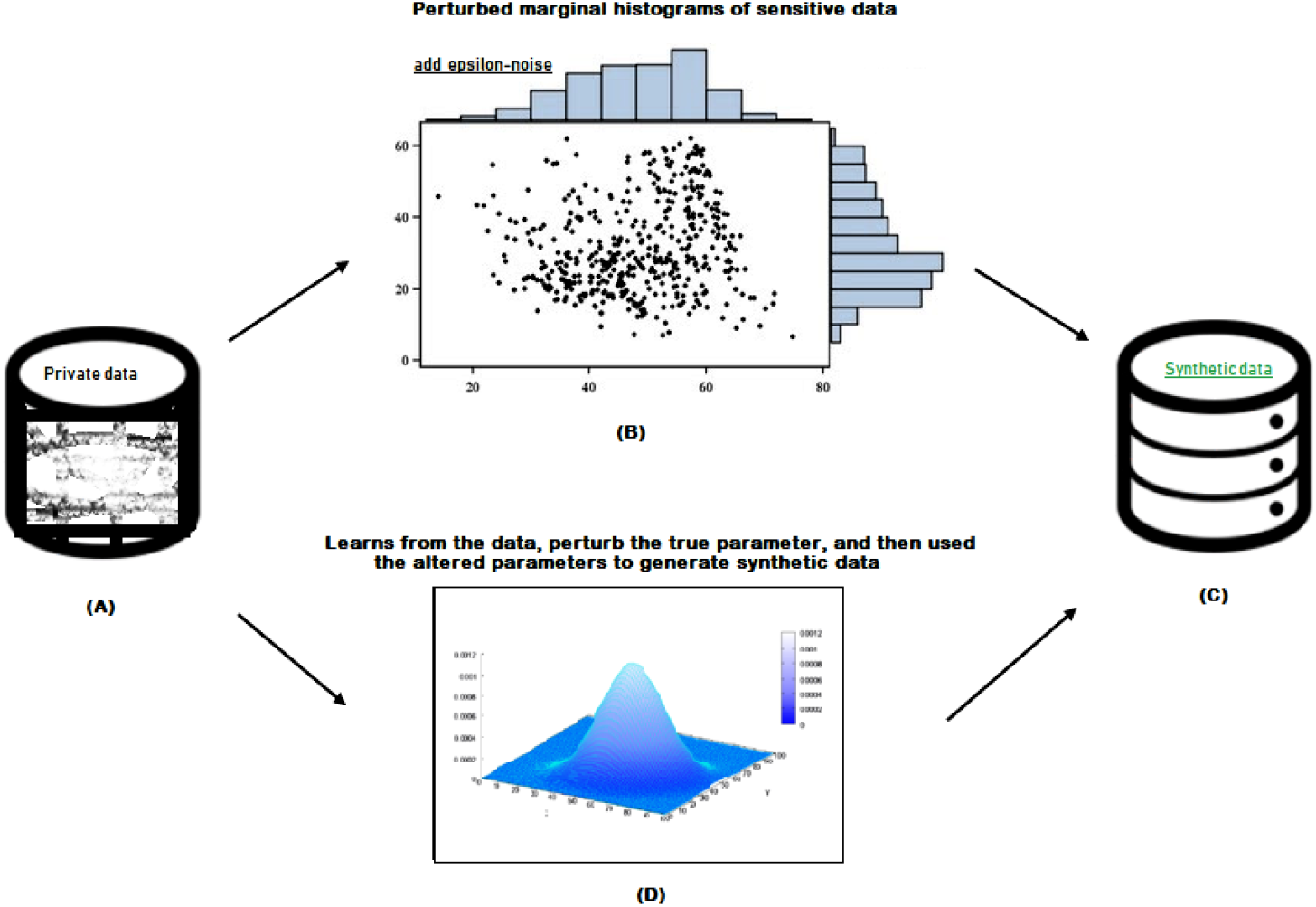
Synthetic data generating architecture

**Figure 2:**
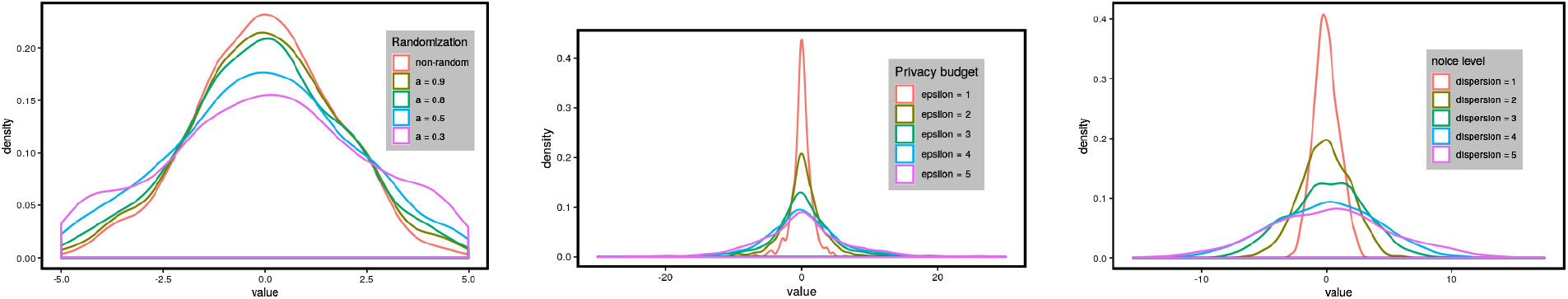
Differential privacy generating mechanisms for different privacy budgets. [L] Randomized response mechanism, [M] Laplace noise mechanism, [R] Exponential noise.

These mechanisms facilitate a range of applications from academic research to industry analytics, ensuring that data privacy does not hinder the potential for data-driven insights.

#### 2.1.2 Pooled synthetic data generation

Pooling promises data privacy with minimal loss of data utility. The method, however, doesn’t provide a formal privacy guarantee nor does it provide a mechanism for quantifying privacy loss when generating synthetic data with the pooling mechanism. More precisely, pooling doesn’t proclaim a guarantee that an individual’s data could not be inferred by an adversary. The cost-effectiveness of pooled analysis and the individual data mask it provides has made it appealing among statisticians and epidemiologists who are able to share aggregate data instead of individual-level data. [8, 6, 9]

Various methods have been proposed for pooling based on a relevant outcome of interest or exposure group. We present below a generic algorithm for generating pooled synthetic data.

##### Algorithm 3

Pooled Synthetic Data Generation

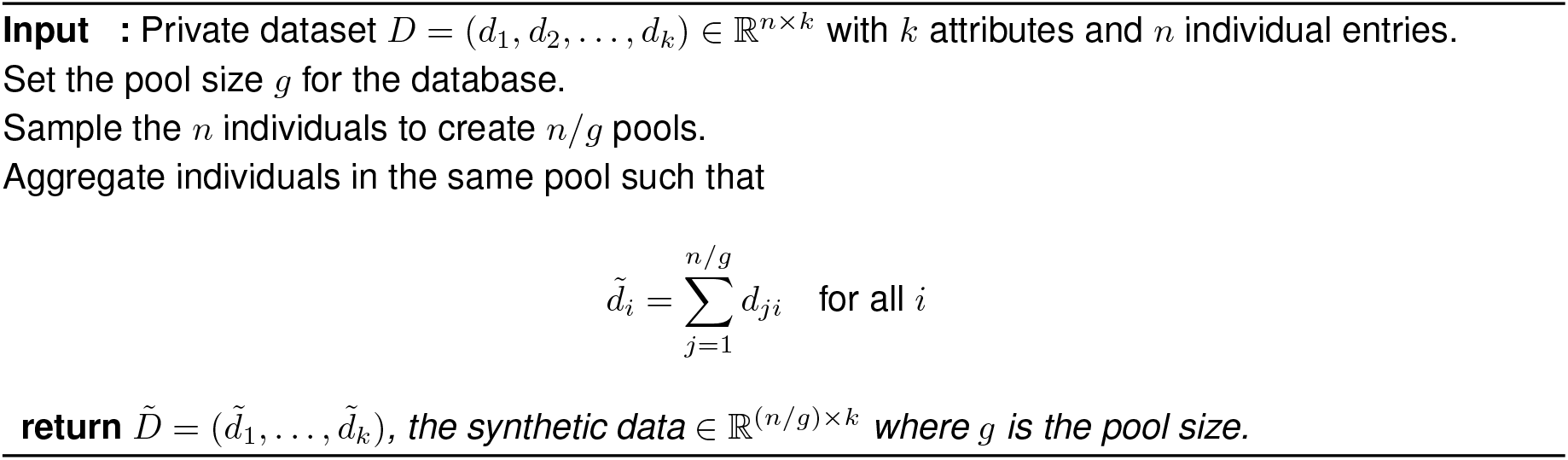

## 3 Pooled (*ϵ, δ*)-Differentially Private Data: PoolDiv

In this section, we introduce PoolDiv, a novel hybrid algorithm that synergistically combines the robust privacy measures of differential privacy from Section 2.1 with the utility-enhancing features of data pooling as described in the Appendix. This approach moderates the stringent privacy parameters typically associated with differential privacy to adopt a more flexible (*ϵ, δ*)-differential privacy model. This relaxation allows for a practical balance between privacy protection and data utility, paving the way for more effective data analysis in sensitive domains.

The main innovation in PoolDiv lies in its two-stage process where differentially private data are first generated with a relaxed (*ϵ, δ*) privacy budget, and subsequently, the data are pooled according to predefined group sizes. This method not only enhances privacy but also reduces computational complexity by minimizing the overhead associated with strict privacy controls. The following algorithm describes the detailed steps involved in generating pooled differentially private data using PoolDiv:

### Algorithm 4

Generation of Pooled (*ϵ, δ*)-Differentially Private Data using PoolDiv

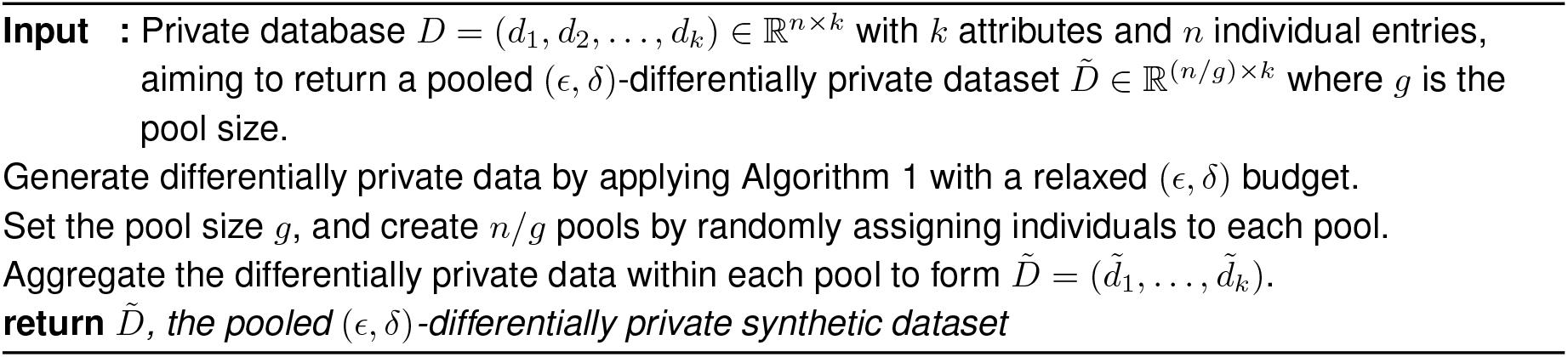

PoolDiv ensures privacy by making the information about any individual indistinguishable from others within the same pool. This is achieved through the differential privacy guarantees applied prior to pooling, which are quantified by the relaxed (*ϵ, δ*) parameters. These parameters are chosen based on the desired level of privacy and the specific requirements of the application context. By adjusting the pooling granularity (i.e., the size of *g*), we can further optimize the balance between data utility and privacy. We hypothesize that PoolDiv, by integrating relaxed differential privacy with pooling, will not only safeguard privacy but also enhance the utility of the synthesized datasets, thereby facilitating more effective and efficient data analysis.

## 4 Regression on Synthetic Databases

Regression analysis is a powerful statistical tool used to model the relationship between a dependent variable and one or more independent variables. In the context of synthetic databases, this technique helps us understand how well the synthetic data can replicate the relationships present in the original data [17] or if bias within the data might have an impact of statistical modeling [18, 19]. Let us consider a regression scenario where *y* = (*y*_1_, …, *y*_*n*_)^*T*^ ∈ ℝ^*n*^ represents the measured responses for *n* individuals, and *X* ∈ ℝ^*n*×*p*^ is the non-random design matrix of predictors. In this matrix, *x*_*i*1_ = 1 for each *i* = 1, …, *n*, to incorporate the intercept term in the regression model.

The model assumes that *y* is a realization of the linear relationship:

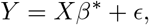

where 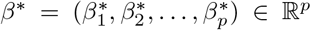 represents the vector of true regression coefficients, and *ϵ* = (*ϵ*_1_, …, *ϵ*_*n*_)^*T*^ is the vector of error terms. These error terms are assumed to be independently and identically distributed (iid) with a mean of zero (E[*ϵ*_*i*_] = 0) and a constant variance 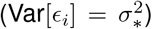. This assumption of homoscedasticity (constant variance) is crucial for the standard least squares estimation to provide the best linear unbiased estimates (BLUE).

To estimate the regression coefficients from the synthetic data, we employ the least squares criterion, which is formulated as follows:

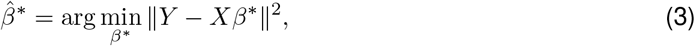

where 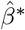 represents the estimated coefficients obtained from minimizing the sum of squared residuals between the observed outcomes and those predicted by the model. This vector of estimates includes 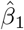, the intercept, and 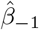, the coefficients associated with the predictors other than the intercept.

In synthetic data analysis, it is essential to compare the estimated coefficients 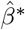 derived from the synthetic data to those obtained from the original data. Such comparisons are crucial for validating the quality and utility of the synthetic data, particularly how well it preserves statistical properties like means, variances, and relationships between variables. Since the intercept can vary significantly with scaling transformations of the dataset, our analysis focuses primarily on 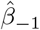, the coefficients of the predictors, which are less sensitive to such transformations. This approach allows for a more stable and meaningful assessment of the synthetic data’s fidelity to the original data.

The performance of the regression model on synthetic data can be further evaluated by computing various goodness-of-fit measures such as the R-squared value, Root Mean Square Error (RMSE), and Mean Absolute Error (MAE). These metrics provide insight into how closely the synthetic data approximates the real data’s underlying structure and variability, thus indicating the practical utility of the synthetic data generation process in preserving key statistical characteristics.

### 4.1 Simulation Study

To evaluate the performance of our proposed mechanisms, we conducted a comprehensive simulation study. We generated three covariates (*X*_1_, *X*_2_, *X*_3_) from Normal distributions, each pair having a correlation coefficient of *ρ*_1,2_ = *ρ*_2,3_ = *ρ*_1,3_ = 0.15. This setup models realistic scenarios where variables are not entirely independent, which is common in many fields such as economics, social sciences, and biostatistics.

We then constructed the outcome variable *y* using a linear additive model:

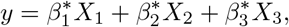

where 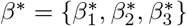 are the true regression coefficients set to known values for the purpose of the simulation. This model helps to understand how well synthetic data can preserve the relationships inherent in the original data when subjected to privacy-preserving algorithms.

The estimates of *β*^*^ using synthetic data generated by our differential privacy mechanisms (histogram perturbation, multinomial-Dirichlet synthesis, and data pooling) are presented in Table 1. We assess how closely these estimates match the true coefficients, which serves as a measure of data utility post-synthesis.

**Table 1:**
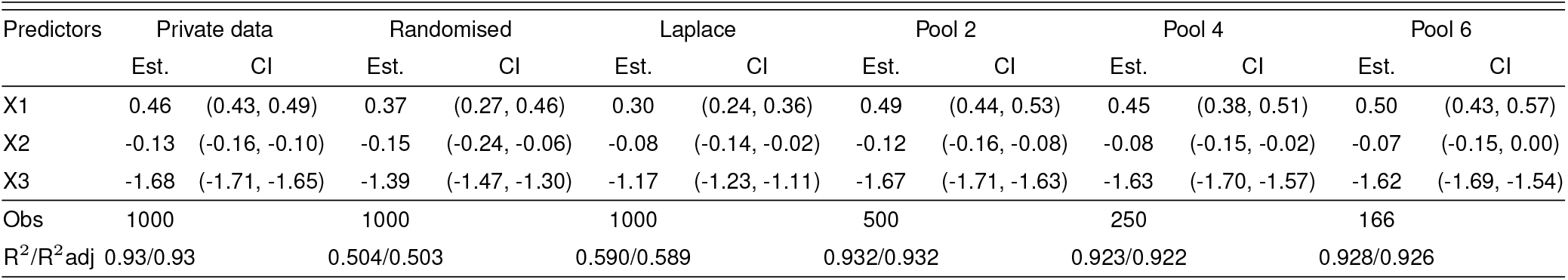
Regression coefficients and confidence intervals for private and synthetic data.

**Table 2:** Regression Estimates Across Different Pooling Divisions.

| Predictors | Pooled-div 2 |  |  | Pooled-div 4 |  |  | Pooled-div 6 |  |  | Pooled-div 10 |  |  |
| --- | --- | --- | --- | --- | --- | --- | --- | --- | --- | --- | --- | --- |
|  | Estimates | CI | p | Estimates | CI | p | Estimates | CI | p | Estimates | CI | p |
| (Intercept) | 0.01 | (-0.17, 0.18) | 0.953 | 0.01 | (-0.34, 0.37) | 0.953 | 0.02 | (-0.52, 0.56) | 0.943 | 0.03 | (-0.86, 0.91) | 0.952 |
| X1 | 0.34 | (0.21, 0.48) | <0.001 | 0.37 | (0.18, 0.55) | <0.001 | 0.24 | (0.02, 0.46) | 0.035 | 0.50 | (0.22, 0.78) | 0.001 |
| X2 | -0.15 | (-0.27, -0.03) | 0.017 | -0.03 | (-0.21, 0.16) | 0.780 | -0.12 | (-0.35, 0.12) | 0.326 | 0.16 | (-0.15, 0.47) | 0.317 |
| X3 | -1.46 | (-1.58, -1.33) | <0.001 | -1.45 | (-1.64, -1.27) | <0.001 | -1.36 | (-1.59, -1.13) | <0.001 | -1.24 | (-1.53, -0.95) | <0.001 |
| Obs. | 500 |  |  | 250 |  |  | 166 |  |  | 100 |  |  |

Following this, we used our hybrid algorithm, referred to as PoolDiv (Algorithm 4), to generate (*ϵ, δ*)-differentially private synthetic data. We experimented with different pool sizes *g* = (2, 4, 6, 8) to explore how the granularity of pooling affects the accuracy of the parameter estimates.

The effectiveness of the regression models fitted to each dataset was quantitatively evaluated using three metrics: Mean Absolute Error (MAE), Root Mean Square Error (RMSE), and R-squared (*R*^2^). These metrics help in understanding different aspects of model accuracy and fit quality: - **MAE** measures the average magnitude of the errors without considering their direction, serving as a clear indicator of average error magnitude. - **RMSE** provides a measure of error magnitude squared, thus giving higher weight to larger errors. This is particularly useful in emphasizing outliers or larger deviations from the mean. - \*\**R*^2^** offers insights into the proportion of variance explained by the model, indicating the strength of the relationship captured by the synthetic data.

#### Interpretation

The results from the simulations provide critical insights: - Models using pooled synthetic data consistently yielded closer estimates to the true parameters, suggesting that pooling might help in mitigating the distortion effects introduced by differential privacy noise. For instance, pooling four observations together (g=4) resulted in unbiased parameter estimates, significantly enhancing model accuracy. - On the contrary, increasing the pool size beyond a certain point seemed to deteriorate the quality of estimates, likely due to over-smoothing or loss of critical data variability. - The performance comparison across various synthetic datasets (as shown in Figure 3) highlights that while differentially private datasets generally perform worse than the original data, the introduction of pooling mechanisms tends to improve performance substantially.

**Figure 3:**
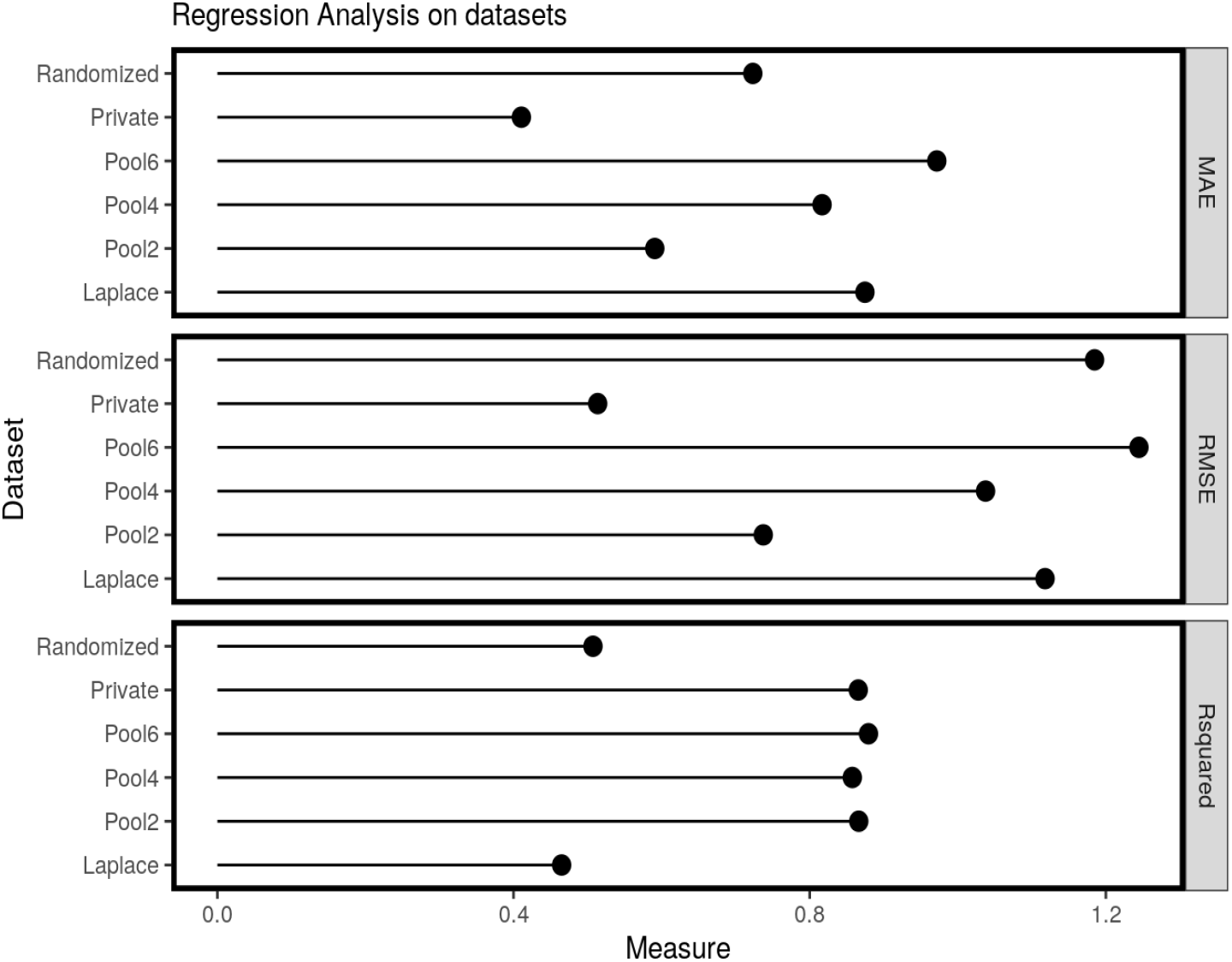
Comparison of model performance across different metrics. RMSE is used as the standard criterion for model comparison.

**Figure 4:**
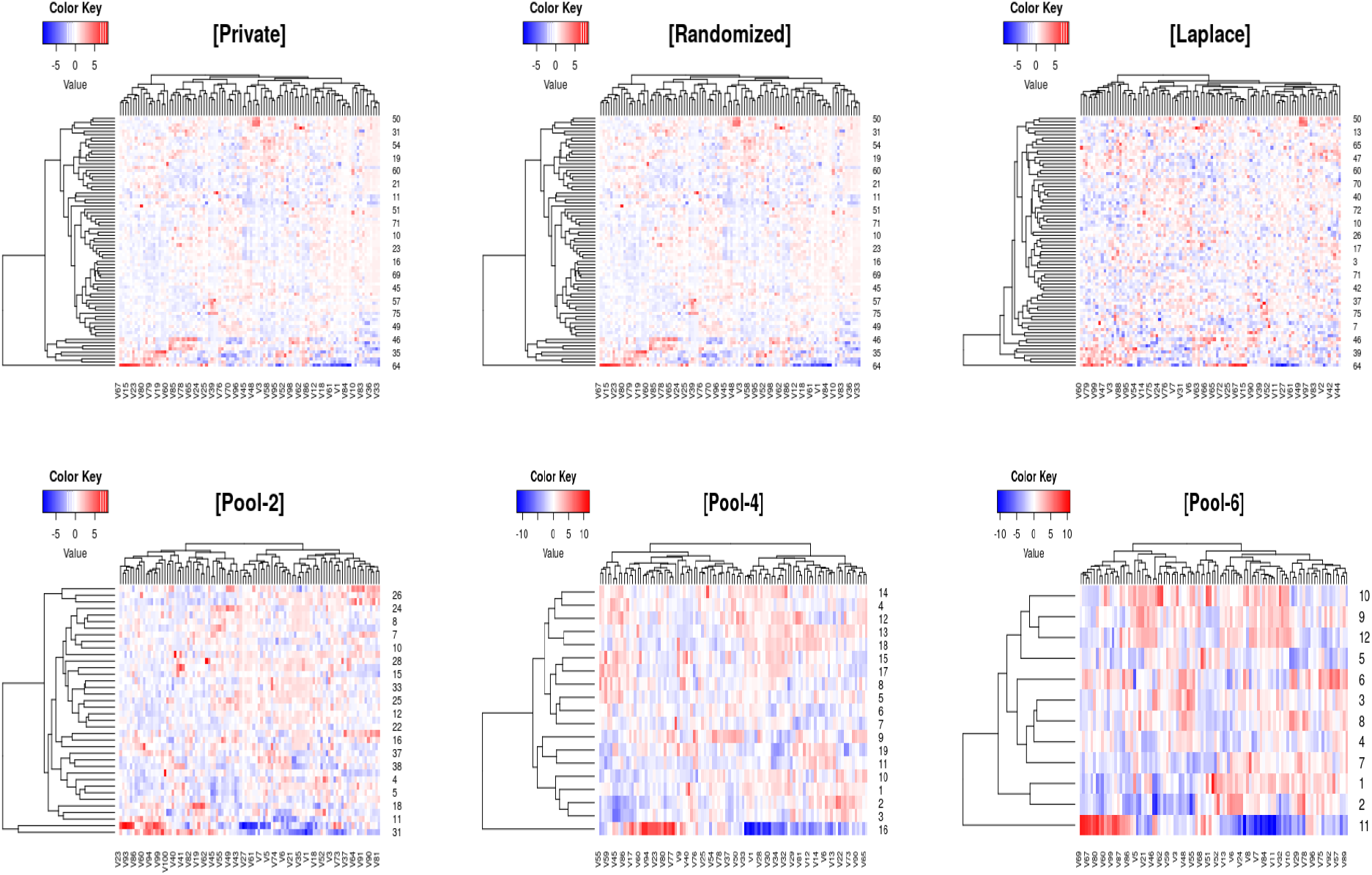
[Private] clustering of the real dataset studied in [20], [Randomized] Synthetic dataset generated via outcome randomization, [Laplace] via laplace noise perturbation of independently generated histograms, [Pool 2,4,6] Synthetic data generated via pooling samples of sizes 2,4,6 respectively

Ultimately, these simulations underscore the importance of choosing appropriate parameters and mechanisms depending on the specific needs of the dataset and the privacy-utility balance required. Our findings suggest that hybrid approaches like PoolDiv can offer a promising compromise, effectively balancing privacy concerns with the need for high-quality synthetic data.

### 4.2 Case Study

The effectiveness of the algorithms was also tested using a real dataset. We analyzed the lymphoma microRNA data reported by Shipp et al. [20]. This dataset comprises a significant subset of participants diagnosed with non-Hodgkin lymphomas (DLBCL), specifically between 30% and 40% of the study’s total population. In total, the study included 58 patients with DLBCL, of whom 32 were successfully cured, while the remaining 26 suffered from fatal or refractory outcomes. The dataset includes measurements of 6,817 gene expression levels. These measurements were used to explore the potential for curative outcomes in patients undergoing a CHOP-based chemotherapy regimen, consisting of cyclophosphamide, adriamycin, vincristine, and prednisone. The analysis aimed to understand how gene expression could influence the effectiveness of this treatment in different patient outcomes.

To assess this high dimension data, we employ a heuristic evaluation of the clustering pattern of the synthetic datasets generated. We present below heatmaps of the synthetic dataset generated. More specifically, we compare the clustering patterns of the private data, randomized outcome data, laplace noise perturbation data, and pooled synthetic datasets.

#### Interpretation

In the heatmaps presented in Figure **??**, we see that the true underlying structure is preserved in the pooled dataset and more prominently exhibited as we continue to pool more observations. In the DP mechanism (e.g. Laplace perturbation), we see completely altered noise profiles. This is unsurprising, as DP has been shown to destroy underlying, local data structure.

## 5 Discussion

Our research has demonstrated that inferential accuracy from traditional differentially private (DP) mechanisms typically falls short compared to pooled analysis. However, our proposed hybrid model, PoolDiv, effectively bridges this gap by combining the robust privacy assurances of DP, specifically under relaxed privacy budgets, with the enhanced utility found in pooled analysis. The performance of the PoolDiv mechanism is on par with traditional DP approaches for low-dimensional data regression and excels in high-dimensional data synthesis.[21, 22, 23]

The PoolDiv mechanism offers significant advantages, particularly in terms of computational efficiency and data structure preservation. Synthetic data generated by PoolDiv tend to be of lower dimensionality, reducing the computational overhead in downstream analyses. For instance, the complexity of estimating regression coefficients in a dataset synthesized by PoolDiv scales as 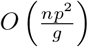, where *g* represents the pool size, thereby reducing computational cost compared to more complex models. Additionally, PoolDiv excels in maintaining the integrity of the underlying data structure, which is crucial for the validity of subsequent analyses.

Simulation results underscore several key insights: Pooling consistently outperforms traditional DP methods in terms of bias reduction and error rates, as evidenced by lower RMSE scores in pooled synthetic data. Moreover, the hybrid PoolDiv mechanism delivers comparable, if not superior, model fits relative to those achieved using randomized algorithms or Laplace noise perturbation techniques.

For high-dimensional data, standard DP techniques often prove inadequate and present significant challenges. The majority of methods capable of generating usable synthetic datasets, such as those based on neural network models (e.g., see [24, 25]), not only require extensive computational resources but also pose steep learning curves for those not versed in advanced machine learning techniques. In contrast, the PoolDiv approach is not only more straightforward to implement but also effectively preserves essential data characteristics, making it highly beneficial for practical applications.

Despite its strengths, the PoolDiv algorithm is not devoid of limitations. The simplistic nature of the outcome-randomized mechanism it employs, while facilitating plausible deniability, may lead to compounded errors when extensive pooling is applied. This is particularly noticeable when more than two samples are pooled, which can skew the results unfavorably. A potential enhancement could involve integrating Laplace noise perturbation for handling high-dimensional data, although this would need to be carefully balanced to avoid exacerbating the computational complexity.

In summary, while the PoolDiv mechanism represents a significant advancement in synthesizing differential privacy-protected data, continuous improvements and adaptations will be essential to address the evolving challenges in data privacy and synthetic data generation.

## Data Availability

All data produced in the present study are available upon reasonable request to the authors.

